# Discrepancies in widely used serological methods to detect *Borrelia* infections may cause missed Lyme diagnoses

**DOI:** 10.1101/2025.03.24.25323417

**Authors:** Elena Volokhina, Annette Stemerding, Yesper Smits, Jordi Lankhof, Milou Kouwijzer, Anja Garritsen

## Abstract

Accurate diagnosis of Lyme disease, caused by *Borrelia burgdorferi*, is essential for timely treatment and prevention of long-term complications. This study investigates discrepancies in widely used serological assays and their impact on diagnostic sensitivity in high-risk populations. We analyzed samples from an annual Lyme screening program conducted by Innatoss Laboratories between November 2020 and May 2022, targeting individuals with occupational exposure to ticks. A retrospective cohort of 60 positive serum samples from 57 participants (aged 23–64) was evaluated using three tier one protocols: EUROIMMUN/ZEUS ELISA, SERION ELISA, and Diasorin CLIA, followed by Viramed immunoblots for confirmation.

Our findings revealed significant variability in test sensitivity: the EUROIMMUN/ZEUS protocol detected 96.8% of cases, compared to 84.1% for SERION (p = 0.023) and 69.8.0% for Diasorin (p<0.001). Notably, six recent *Borrelia* infections were missed due to test selection, as confirmed by longitudinal comparison with previous samples. One case study illustrated the clinical consequences of missed diagnoses, including delayed treatment and symptom persistence.

These results underscore the need for standardized serological protocols and routine longitudinal sample comparison. We propose the establishment of a consensus panel to define assay performance criteria and recommend integration of electronic health records with laboratory systems to enable automated historical data retrieval. Adoption of more sensitive testing strategies and harmonized interpretation guidelines could significantly improve diagnostic accuracy and patient outcomes in Lyme disease.

**Summary box:** *What is already known about this subject:*

- Lyme disease diagnosis relies heavily on serological testing.
- The two-tier testing protocol is widely used but varies in sensitivity depending on the assays employed.
- Serological tests cannot distinguish between past and recent infections without longitudinal data.
- High-risk populations, such as outdoor workers, benefit from regular Lyme screening to detect missed infections.

*What are the new findings:*

- Significant discrepancies that exist between commonly used serological assays affect diagnostic outcomes.
- Six recent *Borrelia* infections were missed due to the choice of test.
- Comparing current results with previous samples from the same individual improves diagnostic accuracy.

*What is the possible impact of this study on clinical practice in the foreseeable future?:*

- Increased awareness of test limitations may lead to earlier and more accurate treatment, reducing long-term complications.
- Standardization and harmonization of Lyme serology can improve future diagnostic guidelines and laboratory practices.
- Routine comparison of longitudinal samples should become standard practice in Lyme screening programs.

## Introduction

Lyme disease is caused by bacteria of the *Borrelia burgdorferi sensu lato* species. Most cases in Europe are caused by *B. afzelii* or *B. garinii*, but infections with *B. burgdorferi sensu stricto*, *B. bavariensis, B. spielmanii, and B. lusitaniae* are also described. In the USA and Canada most Lyme cases are caused by *B. burgdorferi sensu stricto* with some infections by *B. mayonii* (1, 2).

Antibodies against *Borrelia* in Europe are more prevalent in groups with higher exposure to ticks than in the general population (3), which underscores higher rates of *Borrelia* infection in these groups. Moreover, Lyme disease is recognized by the Dutch government as an occupational hazard for outdoor workers in forestry, gardening, landscape and wild-life management, agriculture and other areas (4).

Diagnosis of Lyme disease relies on the history of tick bites and clinical symptoms. However, missed *Borrelia* infections occur frequently. Only 25-73% of Lyme disease patients recall having a tick bite (5). Erythema migrans (EM), the earliest symptom of Lyme borreliosis, is not noticed in approximately 20% of cases, which therefore remain untreated and may progress to Lyme arthritis (60%), Lyme neuroborreliosis (15%) and Lyme carditis (5%); also ocular involvement is frequently described (2, 5–9). To protect high-risk groups, regular screening for *Borrelia* antibodies allows detection of missed infections, timely start of antibiotic treatment and prevention of further complications. Innatoss Laboratories performs an annual Lyme screening for individuals with high occupational exposure to ticks. Screening is done preferably in colder months (October–February), when ticks are not or less active.

An important drawback of any serological testing is the inability to distinguish between antibodies produced during recent infection, which still requires treatment, and an already cleared infection in the past. Therefore, in the annual Lyme screening performed by Innatoss, positive results are always compared with the previous measurement of the same subject. An increase in one or more enzyme-linked immunosorbent assay (ELISA) values, supported by changes in an immunoblot is considered a strong indication of a *B. burgdorferi* infection acquired since the previous measurement.

The annual Lyme screening is performed according to the two-tier testing algorithm, which consists in tier one of the EUROIMMUN Anti-*Borrelia* plus VlsE ELISA (IgG) and EUROIMMUN Anti-*Borrelia* ELISA (IgM). Additionally, the ZEUS *Borrelia* VlsE1/pepC10 IgG/IgM ELISA is used (8, 10). The ZEUS test contains VlsE and contributes to recognition of this important antigen (11–13). If one or more ELISA tests in tier one is positive, *Borrelia* ViraStripe® IgG and/or IgM immunoblot by Viramed Biotech AG is performed to confirm the presence of antibodies. The immunoblots detect antibodies that appear in both early stage (VlsE), as well as late stage (p83/100) Lyme disease.

There are multiple CE-marked serological tests for Lyme disease. Recently, three individuals were tested positive for *Borrelia* antibodies in the annual Lyme screening, while found negative or inconclusive in other laboratories that use different Lyme tests. To investigate the possible cause of discrepancy, the performance of these tests was compared in the samples of these subjects and in a larger cohort.

## Materials and methods

### Study population and sample collection

This study was conducted within the framework of an annual Lyme disease screening program organized by Innatoss Laboratories, targeting individuals with high occupational exposure to ticks, such as outdoor workers in forestry, landscaping, and agriculture. The study was triggered by the tier one test discrepancies in three initial cases, after which the samples of retrospective cohort were analyzed. Blood samples were collected by venipuncture and processed to serum. The participants were selected randomly from those tested positive for *Borrelia* antibodies during the annual Lyme screening. The sampling was done to match gender, age and geographic distribution of the participants in the annual Lyme screening.

### Serological testing protocols

Serological testing followed a two-tier protocol. Tier one included the following assays:

- Anti-*Borrelia* plus VlsE IgG and Anti-*Borrelia* IgM ELISAs (EUROIMMUN AG) and VlsE1/pepC10 IgG/IgM ELISA (ZEUS Scientific). This approach is used in the annual Lyme screening and is the reference testing in this study.
- LIAISON *Borrelia* IgG and IgM Quant chemiluminescence immunoassays (CLIAs) (DiaSorin S.p.A).
- Classic *Borrelia burgdorferi* IgG and IgM ELISAs (Institut Virion\Serion GmbH).

Samples with positive results in any tier one assay were subjected to tier two confirmatory testing using ViraStripe IgG and/or IgM immunoblots (Viramed Biotech AG). The presence of specific antigen bands (e.g., VlsE, OspC, p41) was used to confirm *Borrelia* antibody positivity. IgG immunoblots with a single positive VlsE antigen band were considered positive. All tests were performed manually according to the respective manufacturers’ instructions. Immunoblots were scanned using CanoScan LiDE 110 scanner (Canon) and analyzed using the ViraScan software (Viramed Biotech AG).

### Longitudinal comparison

To identify recent infections, current serological results were compared with previous samples from the same individuals. An increase in ELISA values, supported by new or intensified bands on immunoblots, was considered indicative of a recent *Borrelia* infection.

### Discrepancy analysis

Three individual cases with discrepant results between laboratories were investigated. Samples were retested using alternative platforms to assess assay sensitivity and diagnostic accuracy. Additionally, a subset of 60 samples from the screening cohort who had had tested positive before were analyzed using all tier one assays and corresponding immunoblots. Sensitivity comparisons were made between the protocols employing EUROIMMUN/ZEUS, SERION, and Diasorin assays. Specificity data provided by the manufacturers in the instruction manual and via personal communication are included.

### Statistical analysis

Sensitivity values were calculated for each protocol based on positive cases identified during the annual Lyme disease screening. Statistical analyses were conducted using Python. Differences between protocols were assessed using the McNemar’s test with Bonferroni correction. P-values less than 0.05 were considered statistically significant. Sensitivity and specificity estimates were reported with 95% confidence intervals (CI).

## Results

### Two initial cases with discrepant results

Two adults (case 1 and case 2) had no detectable *Borrelia* antibodies in the first years that they participated in the annual Lyme screening. In November 2023 (case 1) and January 2024 (case 2), the ELISA values increased, confirmed by the immunoblot (Figure 1). Both participants were referred to a general practitioner (GP). Both GPs repeated testing at different laboratories. In both cases, the IgG and IgM tests ordered by a GP were negative and no antibiotic treatment for Lyme disease was prescribed.

**Figure 1.**
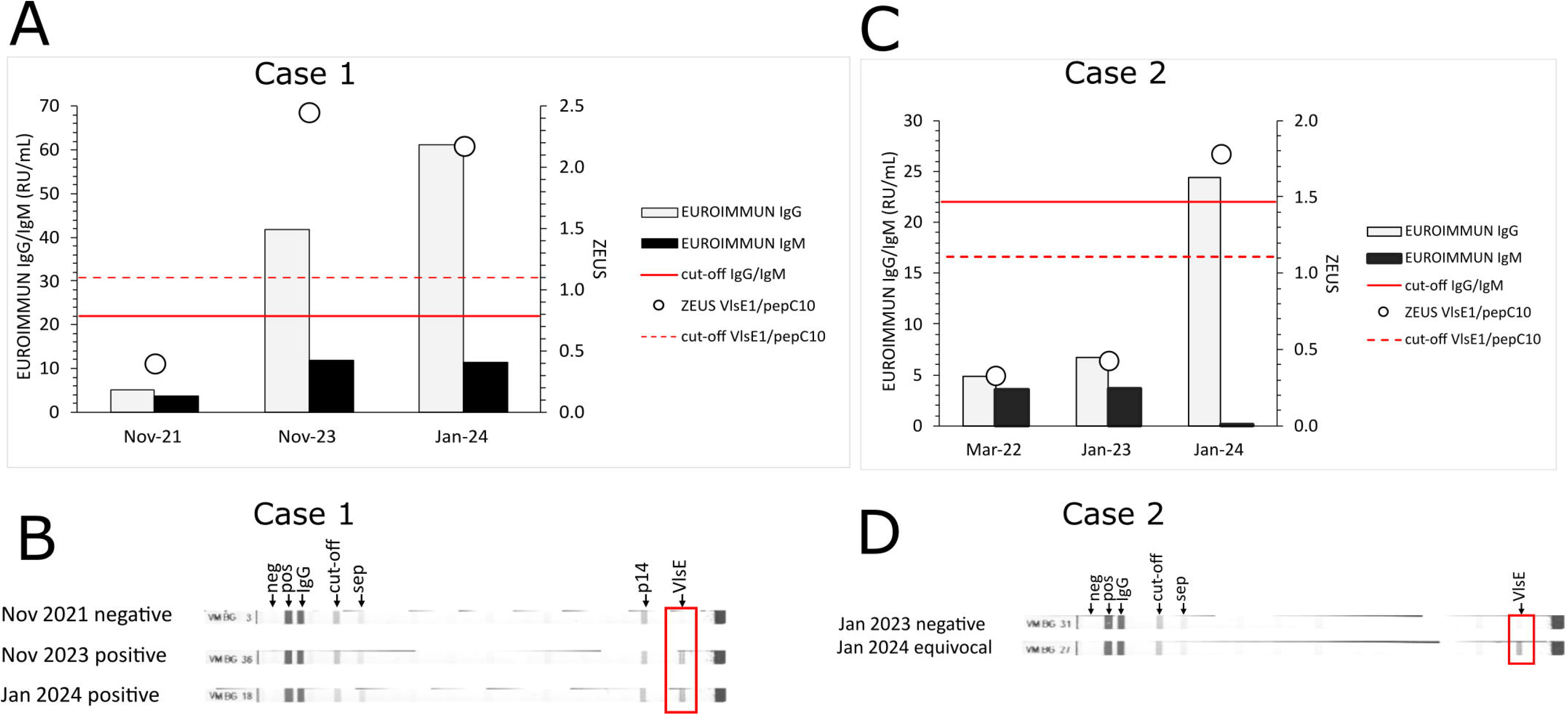
Serological analyses of the serum samples of the two initial cases. Dates of collection are shown as month and year. **A** and **C.** EUROIMMUN IgG and IgM (left Y-axis) and ZEUS VlsE1/pepC10 IgG/IgM (right Y-axis) ELISA results. The cut-offs for positive values are indicated by solid (EUROIMMUN IgG and IgM) and broken (ZEUS VlsE1/pepC10) red lines. RU/mL stands for relative units per milliliter. All results from Diasorin CLIA (case 1) and SERION ELISA (case 2) tests were negative and are not shown. **B** and **D.** Viramed IgG immunoblot results. Positions of negative (neg), positive (pos) and conjugate (IgG) controls; cut-off and separation line (sep) are indicated above the upper strip. Location of the VlsE antigen band is indicated by the red boxes. Overall conclusions per strip are indicated on the left.

In case 1, samples were exchanged with the laboratory that performed the testing for the GP. This laboratory uses Diasorin CLIA for *Borrelia* serology: LIAISON *Borrelia* IgG and LIAISON *Borrelia* IgM Quant. The samples taken in November 2023 and January 2024 showed an increase in EUROIMMUN IgG and ZEUS VlsE1/pepC10 IgG/IgM ELISA of Innatoss (Figure 1A), which was confirmed by the immunoblot (Figure 1B). These samples were negative in the IgG and IgM CLIA of the GP’s laboratory.

In case 2, the analyses ordered by GP were done using SERION IgG and IgM ELISAs. It was not possible to exchange samples for this case. Therefore, the SERION ELISA *classic Borrelia* burgdorferi IgG and IgM tests were implemented at Innatoss to perform the analyses. In the sample from Januari 2024, the SERION IgG and IgM ELISA results were negative, while the EUROIMMUN IgG and ZEUS VlsE1/pepC10 IgG/IgM ELISA showed an increase in antibodies (Figure 1C), which was confirmed by the immunoblot (Figure 1D).

### *Borrelia* infections in a set of samples

To study how often discrepancies between the tests occur and lead to missed Lyme diagnoses, a selection of samples available from the annual Lyme screening cohort was tested. This cohort consists of a high-risk population of over 5000 participants. The prevalence of *Borrelia* antibodies in this group was 15-25% in the period of 2016-2024, while reported prevalence for the general population in the Netherlands between 2016-2017 was only 4.4% (14).

From this retrospective cohort, 60 positive samples from 57 individuals were randomly selected. The individuals were employees of 36 organizations from all 12 Dutch provinces. The cohort consists of 50 males and 7 females between the ages of 23 and 64 years old (median 51), which is typical for this high-risk population. None of the participants reported Lyme symptoms, therefore three controls with clinically confirmed EM (C1-C3) were included. The samples and controls were tested for this study by Innatoss in the SERION IgG and IgM ELISA and by MVZ Dr. Stein + Kollegen (Mönchengladbach) in the Diasorin IgG and IgM CLIA (Table 1). The two-tier protocols that use SERION ELISA and DIASORN CLIA showed lower sensitivity, compared to the EUROIMMUN/ZEUS protocol (Table 2). SERION IgG and IgM tests missed seven samples and Diasorin IgG and IgM CLIA missed 17 samples. Remarkably, one of the symptomatic controls (C2) was missed by the Serion protocol (Table 1), but not by the other two protocols. The specificities reported by the manufacturers for the tier one tests are high and similar between the tests (Table 3).

**Table 1.**
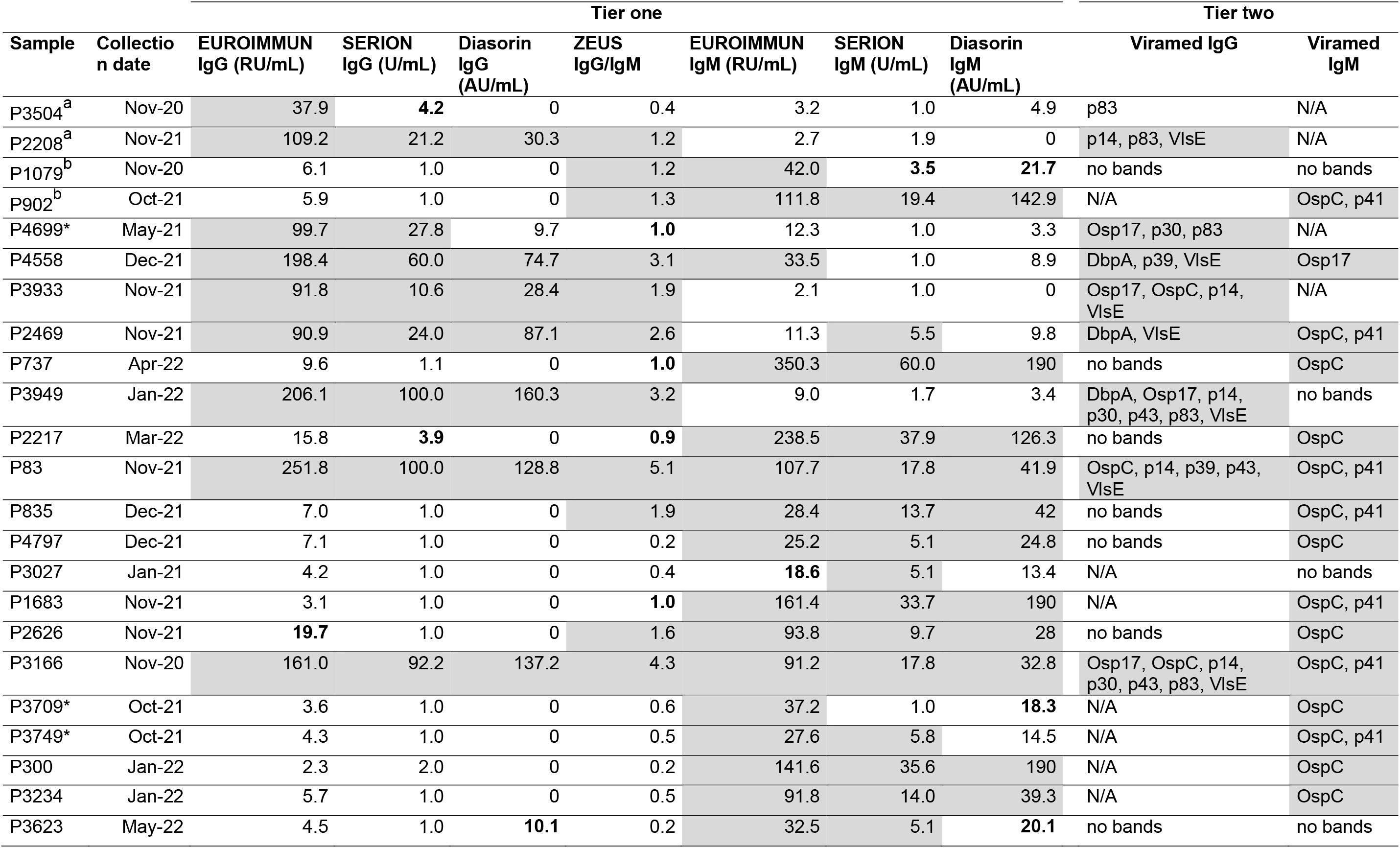
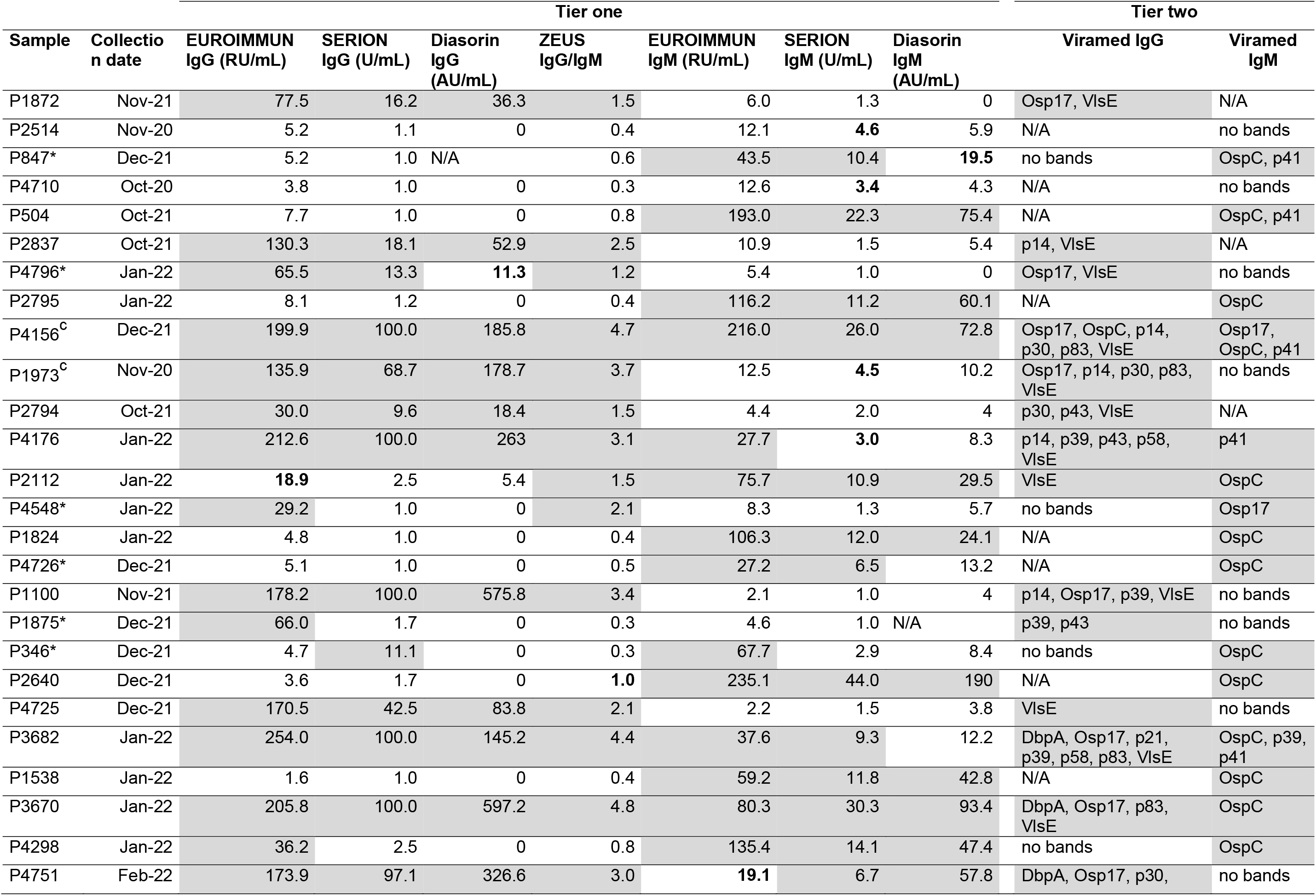
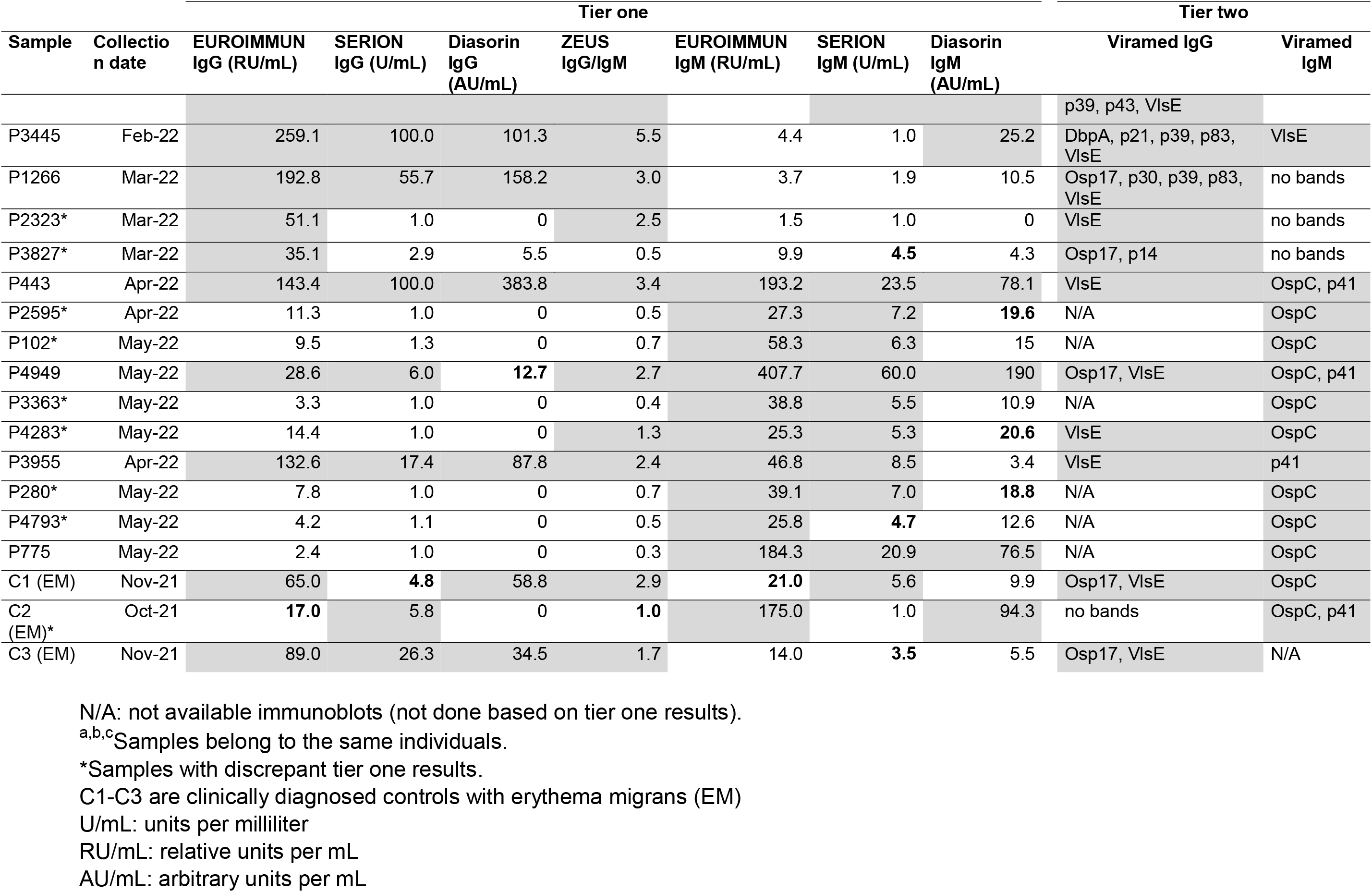
Samples of the participants of the Lyme screening. Diasorin CLIA tests were performed at MVZ Dr. Stein + Kollegen (Mönchengladbach), all other tests were done at Innatoss. Positive results are marked by gray color and equivocal results are shown in bold. Where applicable, units are indicated, the quantitative values are not directly comparable between the test platforms.

**Table 2.**
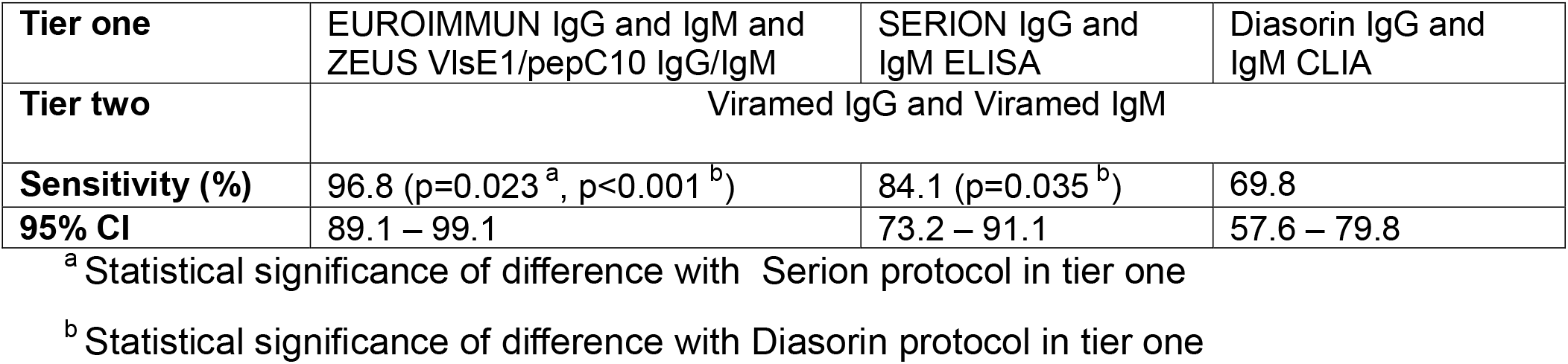
Sensitivity data of the Lyme protocols obtained from analysis of 60 samples and three controls.

**Table 3.** Specificity data of the tier one tests provided by the manufacturers. Total number of samples, mean values (%) and 95% confidence intervals (CI) are indicated.

|  | **EUROIMMUN IgG** | **Serion IgG** | **Diasorin IgG** | **ZEUS** | **EUROIMMUN IgM** | **Serion IgM** | **Diasorin IgM** |
| --- | --- | --- | --- | --- | --- | --- | --- |
| **n** | 92 | 201 | 100 | 400 | 110 | 296 | 91 |
| **Specificity (%)** | 90.2 | 97.0 | 98.0 | 96.5 | 96.4 | 92.6 | 96.7 |
| **95% CI** | 82.4 – 94.8 | 93.6 – 98.6 | 93.0 – 99.7 | 94.2 – 97.9 | 91.0 – 98.6 | 89.0 – 95.0 | 90.8 – 99.1 |

The samples missed by only Diasorin (P3749, P847, P4796 and P4726) and by both Serion and Diasorin (P3709 and P4548) were negative in the previous sampling and thus represent new infections (Figure 2).

**Figure 2.**
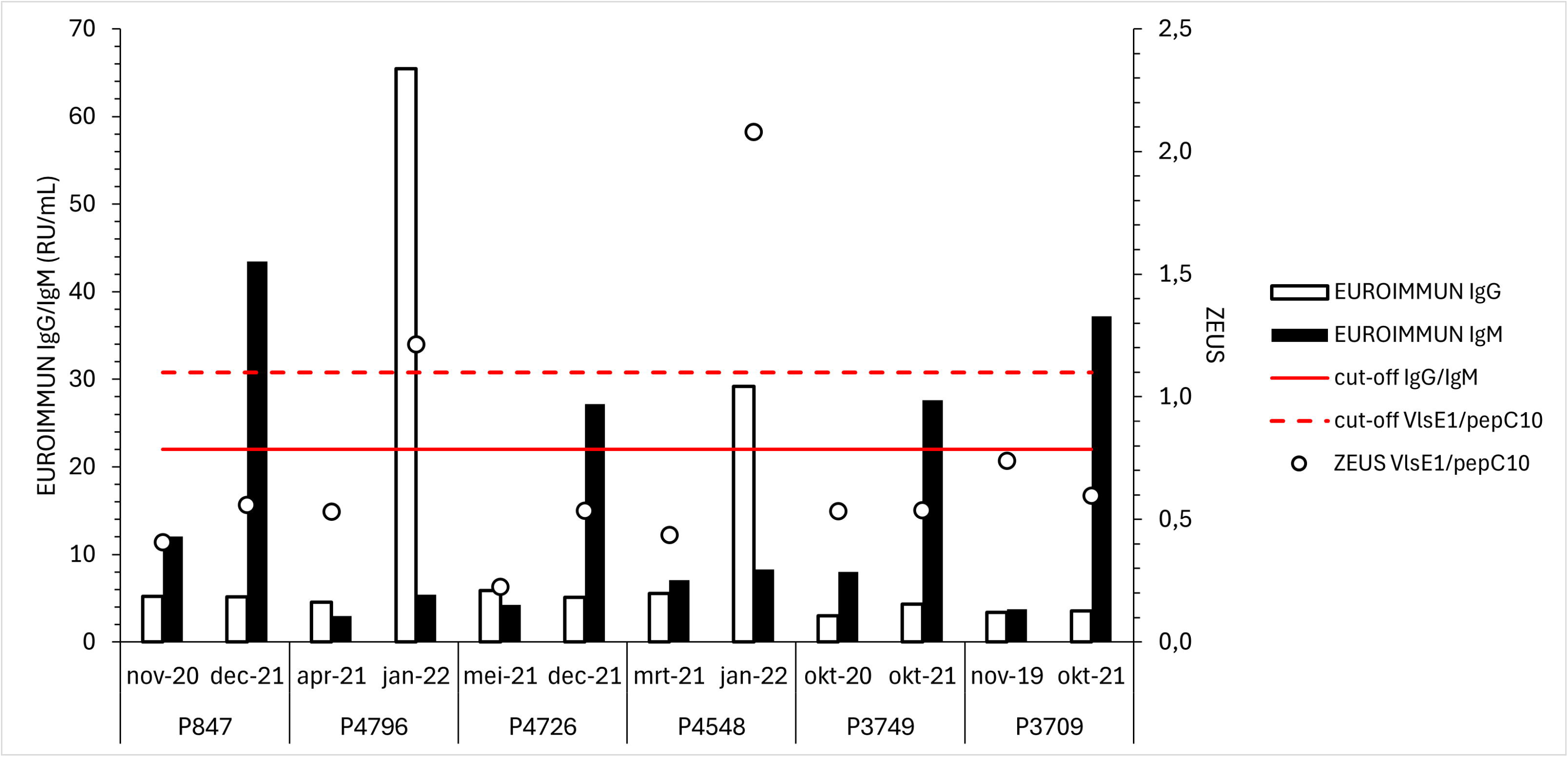
Serological analysis of six subjects that showed an increase in antibodies since the previous measurement in EUROIMMUN IgM, IgG (left Y-axis) or ZEUS VlsE1/pepC10 (right Y-axis) ELISA tests but were negative in Diasorin CLIA or SERION ELISA tests (see Table 1). Dates of collection are shown as month and year. The cut-offs for positive values are indicated by solid (EUROIMMUN IgG and IgM) and broken (ZEUS VlsE1/pepC10) red lines. RU/mL stands for relative units per milliliter. Positive values are confirmed by immunoblot (Table 1).

Test discrepancies have important clinical implications, as they may lead to misdiagnosis of Lyme disease and delayed treatment.

This is illustrated by case 3. A teenage patient with fatigue, joint pain and headache which may be indicative of Lyme borreliosis was referred to the Deventer Hospital. As part of the differential diagnosis, Lyme serology using the Diasorin CLIA as tier one test was performed in two samples taken five weeks apart. In both samples, the IgG CLIA test was negative, and the IgM CLIA test was positive, but not confirmed in the recomLine *Borrelia* IgM immunoblot (Mikrogen Diagnostik). When the same samples were tested at Innatoss, the results were positive in EUROIMMUN IgG and IgM ELISA and ZEUS VlsE1/pepC10 ELISA. Viramed IgG immunoblots were negative, but IgM antibodies were confirmed by a positive OspC antigen band (Figure 3), supporting a diagnosis of Lyme disease. The patient received a doxycycline treatment (200 mg/day, 28 days), after which the symptoms were relieved.

**Figure 3.**
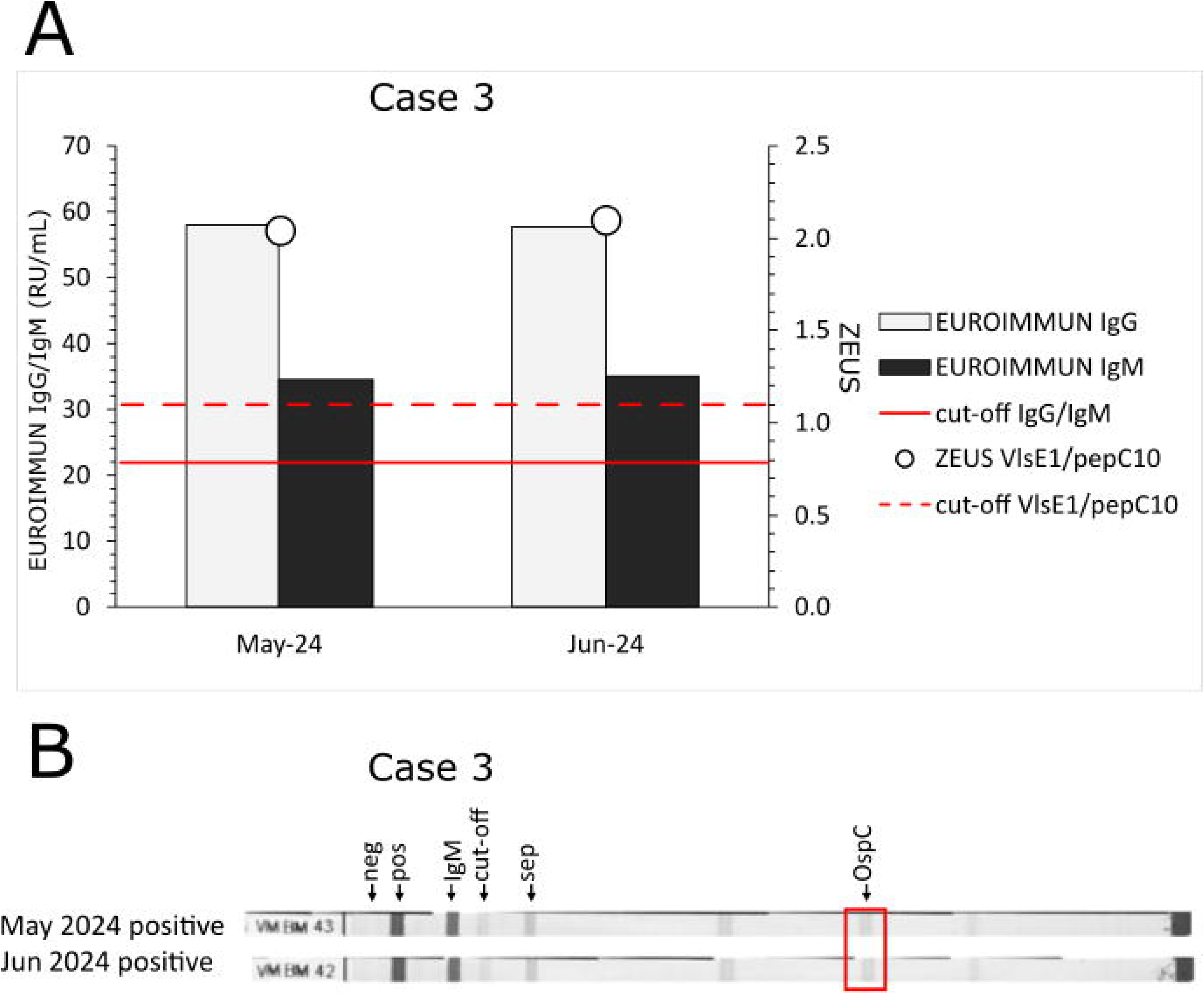
Serological analyses of the serum samples of case 3. Dates of collection are shown as month and year. **A.** EUROIMMUN IgG and IgM (left Y-axis) and ZEUS VlsE1/pepC10 IgG/IgM (right Y-axis) ELISA results. The cut-offs for positive values are indicated by solid (IgG and IgM) and broken (VlsE1/pepC10) red lines. RU/mL stands for relative units per milliliter. Results from CLIA IgM were positive (31 AU/mL (May-24) and 23 AU/mL (Jun-24)) and CLIA IgG were negative for both samples (not shown). **B**. Viramed IgM immunoblot results. Positions of negative (neg), positive (pos) and conjugate (IgM) controls; cut-off and separation line (sep) are indicated above the upper strip. Location of OspC antigen band is indicated by red box. Overall conclusions per strip are indicated on the left. Results of the Mikrogen IgG and IgM immunoblots were negative in both samples (not shown).

## Discussion

The results show that the two-tier Lyme testing, which includes the EUROIMMUN IgG and IgM ELISA and ZEUS *Borrelia* VlsE1/pepC10 IgG/IgM ELISA, detected more positive cases compared to protocols using the Diasorin CLIA and the SERION IgG and IgM ELISA.

The observed discrepancies are consistent with previous reports indicating variability in serological test performance for Lyme disease (15–17). The meta-analysis of 78 studies reported a 50% sensitivity for EM (95% CI: 40% to 61%); 77% for neuroborreliosis (95% CI: 67% to 85%); 97% for acrodermatitis chronica atrophicans (95% CI: 94% to 99%) and 73% for unspecified Lyme borreliosis (95% CI: 53% to 87%) (15). Sensitivity of serological methods is higher in the more advanced stages of the disease. Unfortunately, so are the damaging effects for the patients. Improvement of diagnostics of early borreliosis is therefore extremely important.

Nevertheless, alternatives to the serological testing in Lyme disease are limited and their performances are disappointing. The recent VICTORY study evaluated performance of the three cellular tests for borreliosis and demonstrated that they all have lower specificity than the serological tests, which increases the chance of false-positive outcomes (18). The direct detection of *Borrelia* is challenging due to the low number of bacteria present during infection, Lyme *Borrelia* usually do not cause spirochetemia, except for *B. mayonii* (19). The nucleic acid amplification tests have sensitivity of only 30-50 % in blood of EM patients. When performed on EM biopsies, sensitivity may reach 50-80% however, about 20% of patients do not report EM and thus cannot be diagnosed in this way (2, 5, 8). In the future, direct diagnostics of Lyme disease may be improved by implementing in clinical practice novel techniques such as PCR on *Borrelia* phages or detection of Borrelia-derived peptides using mass spectrometry. However, currently these approaches remain experimental (20, 21).

To date, serological testing is the most reliable method to detect exposure to *Borrelia*, however, choice of the test and testing strategy are important. The discrepancies between different serological tests can lead to missed diagnoses of Lyme disease, as illustrated by the three cases in this study. Misdiagnosis not only may affect a patient’s health and quality of life but also increase healthcare costs due to the need for more intensive treatments in later stages of the disease (6, 7).

Therefore, it is crucial for healthcare providers to be aware of the limitations and strengths of different testing protocols. The use of more sensitive and comprehensive testing methods, such as a broader two-tier protocol including an additional ELISA as used by Innatoss, should be considered to ensure accurate diagnosis and timely treatment.

In 2024, an INSTAND report on external quality assessment for *Borrelia* serology indicated that 42% of the participating laboratories use the Diasorin CLIA; 6% use a SERION ELISA; 26% use the EUROIMMUN ELISAs; data for the ZEUS ELISA are not available (22). Wide-spread use of the Diasorin CLIA can be explained by automation, efficiency and cost-effectiveness of the LIAISON platform, making it attractive, especially for high-volume laboratories. Unfortunately, use of this platform for *Borrelia* serology may lead to missed Lyme diagnoses.

The source of discrepancies between tests is not clear. The lesser sensitivity is likely due to the difference in strains and antigens that are used in the tests (Supplementary table 1). EUROIMMUN and SERION use antigens from different strains to coat wells in the ELISA, in the ZEUS test wells are coated with VlsE1 and pepC10 peptide and the Diasorin test uses recombinant OspC and/or VlsE proteins. However, other factors, such as antigen concentration, sequence and conformation may also affect assay sensitivity. Next to discrepancies in tier one tests, tier two immunoblots may also produce different results, as illustrated in case 3. The difference in sensitivity between immunoblots may also be explained by differences in design of the tests (Supplementary table 1). Multiple antigens in Viramed and Mikrogen blots are different or target different strains (for example OspC and p41 in IgM and IgG immunoblots).

This study has several limitations. The cohort consisted primarily of high-risk individuals undergoing annual screening, potentially limiting applicability to general populations. Additionally, while multiple serological assays were evaluated, not all commercially available tests were included. Finally, the study did not assess long-term clinical outcomes for individuals with missed diagnoses, which could provide further insight into the impact of test sensitivity on disease progression and treatment efficacy.

Future efforts should be directed to further standardization of Lyme serological testing. An international standard, containing relevant antigens should be developed to allow for careful quantification of antibodies in samples, as are available for antibodies to other infectious pathogens, such as *Coxiella burnetii* and SARS-CoV-2 (23, 24).

In addition, interpretation of a single VlsE antigen band on IgG immunoblot should be reevaluated. Currently such blots are interpreted as equivocal, however, if such results are perceived as negative, infections may be missed. This is illustrated by case 2 of this manuscript, where the increase in both the EUROIMMUN IgG and the ZEUS ELISA is accompanied by appearance of only the VlsE antigen band on the immunoblot (Figure 1C and D). In early stages of disease understanding the dynamics of *Borrelia*-specific antibodies is the key to successful use of serology.

Novel approaches may improve Lyme serology by using conserved chimeric antigens, such as BmpA-BBA64 and BmpA-BBK32, to enhance sensitivity and specificity. This strategy could also expand detection to less common human pathogenic genospecies, including *B. bavariensis*, *B. spielmanii,* (25).

In conclusion, our results underscore the importance of serological testing as well as the critical need for standardization and optimization of Lyme disease testing protocols. The use of more comprehensive and sensitive methods, such as the protocol used by Innatoss, should be strongly considered to improve diagnostic accuracy and patient outcomes. Importantly, consensus on the relevance of key antigens such as VlsE should be reached, so interpretation of different immunoblots can be harmonized. To facilitate the adoption of standardized testing protocols, we propose the development of a consensus panel involving clinical microbiologists, infectious disease specialists, and laboratory professionals to define minimum performance criteria for *Borrelia* serological assays. This panel could be coordinated through national or European reference laboratories and supported by external quality assessment programs such as INSTAND.

Furthermore, the practice of comparing current results to previous samples from the same individual should be more widely implemented in routine serological screening for Lyme disease of high-risk groups. Routine longitudinal sample comparison can be implemented by integrating electronic health records with laboratory information systems to flag patients with prior *Borrelia* serology. Laboratories could then automatically retrieve and compare historical data during repeat testing. Exchange of data between laboratories could be coordinated by the consensus panel. Additionally, standardized reporting formats should include a section for longitudinal interpretation, supported by training for laboratory staff and clinicians.

## Samples

This study was conducted using surplus diagnostic material collected during routine annual Lyme screening at Innatoss Laboratories. The reuse of these samples was approved by the Medisch Ethische Toetsingscommissie Brabant, Tilburg, the Netherlands (approval number NW2020-77), which waived the need for additional IRB approval. All participants were informed about the potential use of surplus samples to evaluate diagnostic methods and were given the opportunity to object. Only samples from individuals who did not object were included. Data were handled ensuring confidentiality, secure storage, and restricted access. Only de-identified data were used for analysis, and no personal identifiers were included in the study dataset.

## Data Availability

All data produced in the present study are available upon reasonable request to the authors

## Acknowledgements

We thank Mats van Kempen for technical support and Willemijn Anker and Saskia Theunisse for organizational support for this work.

