## Supplementary table 1 for "Discrepancies in widely used serological methods to detect *Borrelia* infections may cause missed Lyme diagnoses"

**Supplementary Table 1.** Serological tests that are used in this manuscript. Strain information is given as specified by the manufacturer.

| **Assay** | **Method** | **Antigens** |
| --- | --- | --- |
| EUROIMMUN IgG | ELISA | Whole cell extract (Bbss. Ba. Bg) and recombinant VlsE (Bbss) |
| SERION IgG | ELISA | Whole cell extract (Ba. Bg) and recombinant VlsE (Bg) |
| Diasorin IgG | CLIA | Recombinant VlsE* |
| EUROIMMUN IgM | ELISA | Whole cell extract (Bbss. Ba. Bg) |
| SERION IgM | ELISA | Whole cell extract (Ba.Bg) |
| Diasorin IgM | CLIA | Recombinant OspC and VlsE* |
| ZEUS IgG/IgM | ELISA | Recombinant VlsE1. pepC10 (peptide of OspC)* |
| Viramed IgG | Immunoblot | Purified native: OspC (Bbss). p22 (Bbss). Osp17 (Ba). DbpA (Bbss). p14 (Ba)  Recombinant: p83 (Ba. Bbss). p58 (Bbss). p43 (Ba). p39 (Bbss). p30 (Bbss). VlsE (Ba. Bbss. Bbav. Bg. Bs) |
| Mikrogen IgG | Immunoblot | Recombinant: OspC (Bbss. Ba. Bg. Bs). p41 (Bbss). p100 (Ba). p58 (Bg). p39 (Ba). OspA (Ba). p18 (Bbss. Ba. Bg. Bs. Bbav). VIsE (different genospecies) |
| Viramed IgM | Immunoblot | Purified native: OspC (Bbss). p41(Bbss)  Recombinant: p39 (Bbss). Osp17 (Ba). VlsE (Bbss. Ba. Bg) |
| Mikrogen IgM | Immunoblot | Same as Mikrogen IgG |

*Strain of sequence origin is not specified

Bbss (*B. burgdorferi sensu stricto*). Ba (*B. afzelii*). Bg (*B. garinii*). Bbav (*B. bavariensis*). Bs (B*. spielmanii*)
